## Supplementary material for "Ranked severe maternal morbidity index for population-level surveillance at delivery hospitalization based on hospital discharge data": S1. Appendix

**S1 Appendix. The signal to noise ratio: formula for calculation.**

We used the following formula to calculate the signal to noise ratio:

$$R_{j|k}=\frac{h_{j|k}}{\sqrt{{\mu_{j|k}-C}_{k-1}}-\sqrt{C_{k-1}}}$$

Where *h­_j|k_* is the PAF for indicator *j* calculated at iteration *k*, i.e., after recoding the indicator conditions among observations having one of the first *k-1* conditions), *µ_j|k_* is the prevalence of indicator *j* in the study population (after recoding the indicator conditions among observations having one of the first *k-1* indicators), and *C_k-1_* is the cumulative prevalence of all indicators selected before the *k*th step. For the first step, *h­_j|1_* is the ordinary PAF, and *C_k-1_* is zero.

We then selected the indicator with the largest signal-to-noise ratio *R_j|k_* as the *kth* indicator in our list. The value of *h­_j|k_* for the selected indicator is defined to be the hierarchical PAF (hPAF) for the selected indicator. After each iteration, the cumulative hPAF was calculated as the sum of hPAFs for all indicators already selected. The cumulative hPAF after *k* iterations represents the proportion of in-hospital deaths that would be identified using a SMM index comprised of the first *k* indicators selected into the ranked index. Adding hPAFs was possible as our iterative process recodes the indicators to be mutually exclusive.
