## Supplementary material for "Ranked severe maternal morbidity index for population-level surveillance at delivery hospitalization based on hospital discharge data": S1. Table

**S1 Table. Characteristics of study population, Nationwide Inpatient Sample, 1993–2015 (weighted sample = 87,864,173, unweighted sample = 18,198,934).**

|  | **n** | **% (95% CI)** |
| --- | --- | --- |
| Total | 87,864,173 | 100.0 |
| **Age** |  |  |
| 12–18 | 5,526,149 | 6.3 (6.1, 6.5) |
| 19–25 | 28,963,473 | 33.0 (32.5, 33.4) |
| 26–35 | 43,843,239 | 49.9 (49.5, 50.3) |
| 36–45 | 9,464,292 | 10.8 (10.5, 11.0) |
| 46–55 | 67,022 | 0.1 (0.1, 0.1) |
| **Race and Ethnicity** |  |  |
| White, non-Hispanic | 38,434,510 | 43.7 (42.5, 45.0) |
| Black, non-Hispanic | 10,078,680 | 11.5 (10.8, 12.1) |
| Hispanic | 14,779,645 | 16.8 (15.8, 17.8) |
| Other race or ethnicity | 6,650,615 | 7.6 (7.1, 8.0) |
| Missing or unknown | 17,920,723 | 20.4 (18.9, 21.9) |
| **Payer** |  |  |
| Public | 35,172,713 | 40.0 (39.0, 41.0) |
| Private | 46,853,838 | 53.3 (52.2, 54.5) |
| Other | 5,593,060 | 6.4 (5.9, 6.8) |
| Missing or unknown | 244,563 | 0.3 (0.2, 0.3) |
| **Mode of Delivery** |  |  |
| Repeat cesarean | 10,325,192 | 11.8 (11.6, 11.9) |
| Primary cesarean | 14,216,259 | 16.2 (16.0, 16.4) |
| Vaginal | 63,322,722 | 72.1 (71.8, 72.4) |
| **Hospital Region** |  |  |
| Northeast | 14,783,059 | 16.8 (15.6, 18.0) |
| Midwest | 19,396,116 | 22.1 (20.8, 23.4) |
| South | 31,997,346 | 36.4 (34.6, 38.3) |
| West | 21,687,652 | 24.7 (23.2, 26.2) |
| **Hospital Location** |  |  |
| Rural | 10,607,929 | 12.1 (11.3, 12.9) |
| Urban non-teaching | 35,037,086 | 39.9 (38.2, 41.6) |
| Urban teaching | 41,900,794 | 47.7 (45.9, 49.5) |
| Missing or unknown | 318,364 | 0.4 (0.1, 0.6) |
| **Year** |  |  |
| 1993 | 3,711,994 | 4.2 (3.8, 4.7) |
| 1994 | 3,697,459 | 4.2 (3.8, 4.6) |
| 1995 | 3,723,862 | 4.2 (3.8, 4.6) |
| 1996 | 3,677,018 | 4.2 (3.8, 4.6) |
| 1997 | 3,667,243 | 4.2 (3.8, 4.6) |
| 1998 | 3,660,795 | 4.2 (3.8, 4.6) |
| 1999 | 3,741,842 | 4.3 (3.8, 4.7) |
| 2000 | 3,960,075 | 4.5 (4.0, 5.0) |
| 2001 | 3,876,405 | 4.4 (4.0, 4.9) |
| 2002 | 4,002,465 | 4.6 (4.1, 5.0) |
| 2003 | 3,939,389 | 4.5 (4.0, 4.9) |
| 2004 | 4,095,133 | 4.7 (4.2, 5.1) |
| 2005 | 4,058,251 | 4.6 (4.1, 5.1) |
| 2006 | 4,132,023 | 4.7 (4.2, 5.2) |
| 2007 | 4,379,065 | 5.0 (4.5, 5.5) |
| 2008 | 4,046,357 | 4.6 (4.1, 5.1) |
| 2009 | 3,941,419 | 4.5 (4.0, 5.0) |
| 2010 | 3,714,024 | 4.2 (3.8, 4.7) |
| 2011 | 3,670,999 | 4.2 (3.7, 4.7) |
| 2012 | 3,766,201 | 4.3 (4.0, 4.6) |
| 2013 | 3,741,613 | 4.3 (4.0, 4.5) |
| 2014 | 3,801,565 | 4.3 (4.1, 4.6) |
| 2015 | 2,858,975 | 3.3 (3.1, 3.5) |
